# Effectiveness of Typhoid Conjugate Vaccine in Zimbabwe used in response to an outbreak among children and young adults: a matched case control study

**DOI:** 10.1101/2022.03.28.22273032

**Authors:** Maria S. Lightowler, Portia Manangazira, Fabienne Nackers, Michel Van Herp, Isaac Phiri, Kuziwa Kuwenyi, Isabella Panunzi, Daniela Garone, Farayi Marume, Andrew Tarupiwa, Eva Ferreras, Clemence Duri, Francisco J. Luquero

## Abstract

**Background:** Zimbabwe suffers from regular outbreaks of typhoid fever (TF), worse since 2017. Most cases were in Harare and a vaccination campaign with Typhoid Conjugate Vaccine (TCV) was conducted in March 2019. The vaccine effectiveness (VE) was assessed against culture-confirmed *S*. Typhi in children six months to 15 years and in individuals six months to 45 years in Harare.

**Methods:** A matched case-control study was conducted in three urban suburbs of Harare targeted by the TCV vaccination campaign. Suspected TF cases were enrolled prospectively in four health facilities and were matched to facility (1:1) and community (1:5) controls.

**Findings:** Of 504 suspected cases from July 2019 to March 2020, 148 laboratory-confirmed TF cases and 153 controls confirmed-negative were identified. One hundred and five (47 aged six months to 15 years) cases were age, sex, and residence matched with 105 facility-based controls while 96 cases were matched 1:5 by age, sex, and immediate-neighbour with 229 community controls.

The adjusted VE against confirmed TF was 75% (95%CI: 1–94, p=0.049) compared to facility controls, and 84% (95%CI: 57–94, p<0.001) compared to community controls in individuals six months to 15 years. The adjusted VE against confirmed TF was 46% (95%CI: 26–77, p=0.153) compared to facility controls, and 67% (95%CI: 35–83, p=0.002) compared to community controls six months to 45 years old.

**Interpretation:** This study confirms that one vaccine dose of TCV is effective to control TF in children between six months and 15 years old in an African setting.

## Introduction

Typhoid fever (TF), an acute systemic infection caused by *Salmonella* Typhi (*S*. Typhi),^1^ remains a large public health problem worldwide, especially in Southern Asia and sub-Saharan Africa, with an estimated 9-12.5 million cases and 65,000-188,000 annual deaths.^2,3^ The TF burden is probably underestimated^4,5^ as enhanced surveillance in endemic Africa and Asian areas has shown high heterogeneity in its geographical distribution.^6,7^ Both urban slums^8,9^ and rural areas with poor water and sanitation conditions are prone to TF transmission, with seasonal patterns. *S*. Typhi can also cause large outbreaks.^10^

Zimbabwe has suffered regular outbreaks of TF. Most of the cases were reported from the capital city, Harare, ^8,11^ especially in the densely populated southwestern suburbs. A large number of cases have been reported from 2010 and seasonal outbreaks have occurred annually from October to March. In October 2017 a new outbreak started in Harare. As of June 28, 2018, a total of 4330 cases were reported to the Ministry of Health and Child Care (MOHCC).^12^ The 0-14 age group accounted for 47% of the cases. The southwestern suburbs, characterized by low socioeconomic status, overcrowding, intermittent water supplies, frequent sewer line breaks and low elevation were the most severely affected. The reported number of deaths (n=5) was low (case fatality risk: 0.1-0.2%),^13^ probably because of good access to health services and availability of antibiotics. However, in Harare, the resistance of *S*. Typhi to ciprofloxacin, the first treatment line in Zimbabwe, increased worryingly from 0% in 2012 to 22% in 2017.^14^

Access to safe water, and adequate sanitation and hygiene remain the mainstay of TF prevention and control. As an important complementary tool for endemic and epidemic disease control, the World Health Organization (WHO) recommends programmatic use of typhoid vaccines, preferentially Typhoid Conjugate Vaccine (TCV). TCV shows strong immunological response for all ages and is suitable for young children. WHO also recommends documenting the field experience and impact of different vaccination strategies, as well as their integration with water and sanitation or other public health interventions.^15^

Typbar-TCV®, manufactured by Bharat Biotech International Limited (BBIL) was licensed in India in 2013 for intramuscular administration of a single dose (0.5 mL) in children aged 6 months and older and prequalified by WHO in 2017; each dose comprises 25 μg of purified Vi-capsular polysaccharide conjugated to tetanus toxoid. It is available in single-dose vials or pre-filled syringes, and 5-dose vials. In the multi-dose formulation each dose also contains 5 mg of 2-phenoxyethanol as preservative. The vaccine has a vaccine vial monitor (VVM30) and the manufacturer-recommended storage temperature is 2–8 °C ^15^.

In 2019 in Harare, the MOHCC conducted a mass vaccination campaign of approximately 320,000 Typbar-TCV® doses targeting high-risk populations to control the outbreak and prevent possible peaks in the following seasons. The most affected suburbs (Mbare, Kuwadzana and Glenview) were covered. Between 25^th^ February to 4^th^ March 2019, one dose of Typbar-TCV® was delivered to individuals of six months to 15 years old excluding pregnant women. Considering the high attack rates observed in individuals 15 to 45 years old in Mbare, the target age group was extended up to 45 years old. The vaccination campaign reached a high coverage of 85% in the target age groups.^16^

We conducted a matched case-control study to assess the effectiveness of TCV used under real life conditions in response to an outbreak. We aimed to measure the effectiveness of TCV against confirmed TF among children six months old to 15 years old. In a secondary analysis, we extended the population to measure the effectiveness among individuals aged six months to 45 years following the vaccination target group used in one of the suburbs.

#### Research in context

##### Evidence before this study

We searched PubMed for reports published before September 30, 2021, with terms “typhoid vaccine” OR “typhoid conjugate vaccine” AND “effectiveness”, “efficacy”, “outbreak response” OR “impact”. We searched for studies that assessed the effectiveness or efficacy against confirmed typhoid fever cases by blood culture and/or serology. We identified seven studies in which results were reported on efficacy or effectiveness on Typar-TCV. The vaccine efficacy was 54.6% in an adult volunteer challenge trial conducted in UK. The vaccine efficacy found in three other studies was 85%: a phase III seroefficacy trial in India (among individuals 2 to 45 years), preliminary results from a phase III clinical trial in Nepal and a cluster randomised trial in Bangladesh (the two latest among children aged nine months to 16 years). Two studies were from Pakistan. One cohort study reported vaccine effectiveness of 95% among children between six months to 10 years and in a matched case control study following a mass vaccination campaign it was 72%. Finally, a phase three randomised control trial in Malawi reported 81% efficacy among children aged nine months to 12 years.

This is the first study, to our knowledge, to report the field effectiveness of TCV after a mass vaccination campaign in response to an outbreak in Africa endemic context. Effectiveness data are limited to children and there are only two studies reporting efficacy that includes adults (the ones from UK and India).

##### Added value of this study

In this matched case-control study, we found that one dose of TCV was effective against blood culture-confirmed S. typhi symptomatic children aged six months to 15-years. The effectiveness was higher among community controls (84%), when compared with facility controls (75%); this could be due to selection bias due to the low sensitivity of blood culture. It was lower when was estimated among individuals aged six months to 45 years (67%), which might be due to longer exposure to prior infections among adults. Zimbabwe is the first African country to use TCV in as part of an outbreak response.

##### Implications of all the available evidence

This study confirms that one vaccine dose of TCV can be an effective tool to control TF; it is the first to assess the use of the vaccine under real programmatic conditions in an African setting. These findings are in line with WHO recommendations and the decision by several countries to introduce TCV into routine vaccination.

## Methods

### Study setting and design

We did a prospective matched control study. The study population comprised residents of three suburbs targeted for vaccination (Mbare, Kuwadzana and Glen View). Cases were recruited in four health care facilities: Beatrice Road Infectious Disease Hospital (BRIDH), the main infectious disease hospital in Harare that serves the population from the three suburbs, and the main public health centre (polyclinic) of each suburb.

Eligible TF suspected cases comprised individuals aged six months to 45 years living in the areas of Glen View, Mbare and Kuwadzana at the time of the TCV vaccination campaign and seeking care at participating health facilities. A suspected TF case was defined as any person who presented with fever (38°C and above) that had lasted for at least three days (self-reported). A probable case was a TF suspected case without *S*. Typhi isolation in a blood culture but with a positive sero-diagnosis (defined as a doubling in the IgG Vi titer one month after admission/onset of fever). A confirmed case was a TF suspected case with a laboratory-confirmed positive blood culture of *S*. Typhi.

Facility controls were selected among TF suspected cases without isolation of *S*. typhi in blood culture and without a positive sero-diagnosis after one month.

Community controls were selected among neighbours residing up to 150 meters from suspected TF cases. Community controls were considered eligible if they were aged six months to 45 years old at the start of the vaccination campaign, lived in one of the three suburbs since the TCV vaccination campaign, and had no personal history of TF confirmed by a clinician.

Suspected TF cases were recruited prospectively. The primary analysis included children aged six months to 15 years. Vaccine effectiveness was evaluated using a case-control test negative design as the main study design. We used the test-negative as the main study design as this approach has been suggested as a valid method to estimated vaccine effectiveness in other vaccines.^17^ Confirmed TF cases were individually matched to facility controls on age, gender, and suburb with the closest enrolled date (1:1 ratio). Using a second study design, five community controls were individually matched, on neighbourhood, age (5-years category) and gender to confirmed and probable TF cases (1:5 ratio). The secondary analyses for both methods included estimates of vaccine effectiveness among the whole population (six month to 45 years eligible for vaccination during the campaign.

A vaccinated individual was a person who reported having received a TCV dose during the 2019 vaccination campaign. Vaccination status was verified with the vaccination card if available or by oral ascertainment otherwise (date, administration by injection, place of vaccination). Individuals with unclear vaccination status (two probable TF cases and five community controls) were excluded from the analyses. A household was considered as a group of individuals living under the same roof and eating from the same cooking pot at least three times a week irrespective of family ties.

### Data collection

All suspected TF cases attending health care facilities were screened for eligibility. Following informed consent, study staff administered a standardised questionnaire. The following information was collected: gender, age, profession, place of residence, socio-economic data, history of signs and symptoms of the current episode, contact with TF cases, type of water and food consumed, use of soap, toilets and latrine and TCV vaccination status (Supplementary material).

### Laboratory procedures

All suspected TF cases were tested at enrolment (blood culture, antimicrobial susceptibility testing, and serology test) and at one month after enrolment (stool culture and serology test).

Blood was collected for culture (10 ml of blood for individuals 5-45 years; one to four ml for children under five years) and serological testing (five ml of blood for participants 5-45 years of age; one to three ml for children aged under five years).

All blood samples were transported to and tested at the National Microbiology Reference Laboratory (NMRL) at ambient temperature. The blood samples for serum extraction were then kept at 2-8°C until sample processing.

Blood samples were screened for *Salmonella enterica* serovar Typhi using standard laboratory protocol for blood culture methods.^18^ Confirmed *S*. Typhi isolates were screened for antibiotic susceptibility using the Kirby Bauer disc diffusion method and results were interpreted based on the 2017 CLSI guidelines.^19^

Stool samples were cultured using standard protocols. Individuals with convalescent carriage and a chronic carriage were defined as shedding of S. Typhi in stools 30 days (+/-five days) or 365 days respectively from enrolment among the confirmed TF cases. Stools were also collected from community controls to estimate the prevalence of asymptomatic carriage in populations with active transmission of *S*. Typhi.

Serum anti-Vi immunoglobulin G (IgG) antibody levels were measured using the commercially available VaccZyme enzyme-linked immunosorbent assay kit (The Binding Site), per the manufacturer’s instructions. The lower limit of assay quantification is 7.4 ELISA units (EU)/mL.

#### Data management and statistical analysis

Questionnaires were completed in KoboCollect software installed on a mobile device. Data were regularly uploaded to a password-controlled secure central server.

Characteristics of TF cases and individually matched controls were compared with univariate conditional logistic regression and likelihood ratio test (LRT). Data sparsity was handled by reducing the number of categories in some variables (occupation, highest level of education achieved, main source of treatment water). Missing data were checked for each variable and individuals with missing values were labelled as “unknown” and excluded from the related analysis.

Conditional logistic regression was used to compare the odds of vaccination in confirmed TF cases and controls, and to calculate the matched odds ratio (OR) in children aged six months to 15 years (primary analyses)). Vaccine Effectiveness (VE) was calculated as 1 - OR x 100%. the TCV VE was estimated against confirmed TF and against confirmed or probable TF. A secondary analysis was carried out by including all individuals aged six months to 45 years.

Multivariable conditional logistic regression was performed to incorporate the effects of potential confounders and effect modifiers. We evaluated the difference in demographics, housing and food and personal hygiene risk factors exposure between cases and controls. Potential confounders that modified the VE by more than 10 percentage points were included in multivariable models for analyses. P-values were derived from LRT. All P values and 95% confidence intervals were two-sided. Statistical significance was set at 0.05.

To estimate the required sample size, we hypothesised a VE of 80% with 50% vaccine coverage among participants aged six months and 15 years. Considering an alpha error-risk of 5%, 10% of missing data and a power of 0.8 (against the null VE=0%), 40 confirmed TF cases and 200 matched community controls aged six months and 15 years old were needed in a 1:5 ratio; and 44 confirmed TF cases and 44 matched facility controls in a 1:1 ratio.

Statistical analyses were performed using STATA version 16.1 (Stata Corp, College Station, TX).

#### Bias-indicator analysis

To detect a possible health seeking behavior bias between vaccinated and unvaccinated individuals possibly affecting the VE estimates, we compared the odds of vaccination between facility controls and community controls (five community controls matched on neighborhood, age and gender, 1:5 ratio).

#### Ethics considerations

The study protocol was approved by the Medical Research Zimbabwe Committee (approval number MRCZ/A/2460) and by the Médecins Sans Frontières Ethics Review Board (approval number 18114).

## Results

The enrolment started in July 2019 in Glen View polyclinic and BRIDH, and in October 2019 in Mbare and Kuwadzana polyclinics. Enrolments were stopped by the end of March 2020 due to the COVID-19 pandemic until July 2020 in Mbare polyclinic and until November 2020 in Glen view polyclinic. The study was closed after completion for the primary objective by April 30, 2021.

Overall, 707 suspected typhoid fever cases were screened for eligibility. Of these, 504 were enrolled (Figure 1) and 148 had confirmed *S*. typhi blood culture. The weekly distribution of eligible patients (suspected and confirmed TF cases) is shown in Figure 2. The number of confirmed TF cases along the period of the study coincides with seasonality of TF in Zimbabwe. The results of the blood culture of patients enrolled are shown in Table 1.

**Table 1.** Blood culture results upon enrolment from 499 eligible TF suspect cases from July 2029 to April 2021, Harare, Zimbabwe.

| Study patients |  |  |
| --- | --- | --- |
| Blood Cultures Results | N | Percent |
| No growth | 347 | 69.5 |
| *Salmonella typhi* * | 148 | 29.7 |
| Non-typhoidal *salmonella* | 3 | 0.6 |
| *Enterococcus faecalis* | 1 | 0.2 |
| **Total** | **499** | **100** |

\*47 ≤15 years old

**Figure 1.**
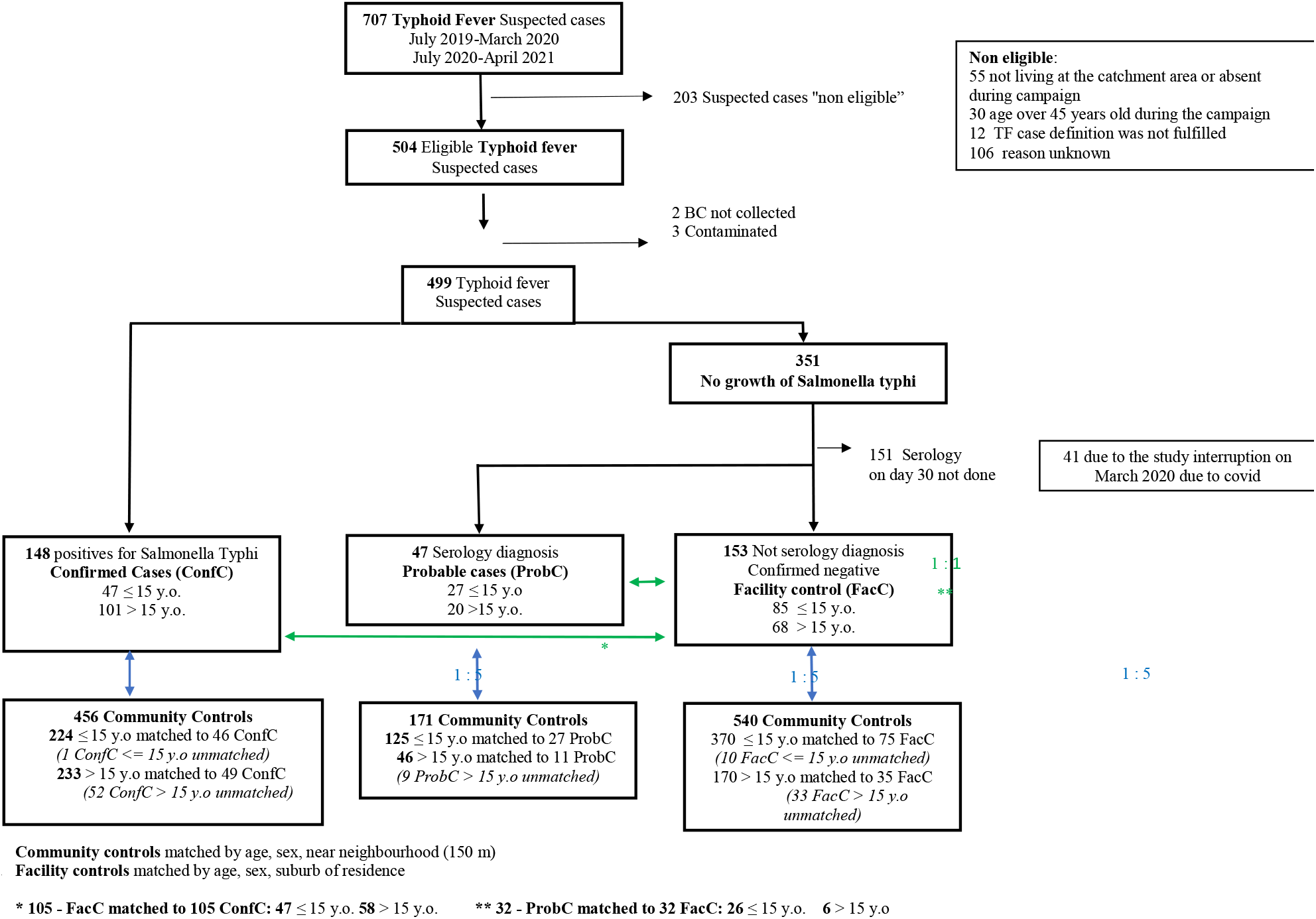

**Figure 2.**
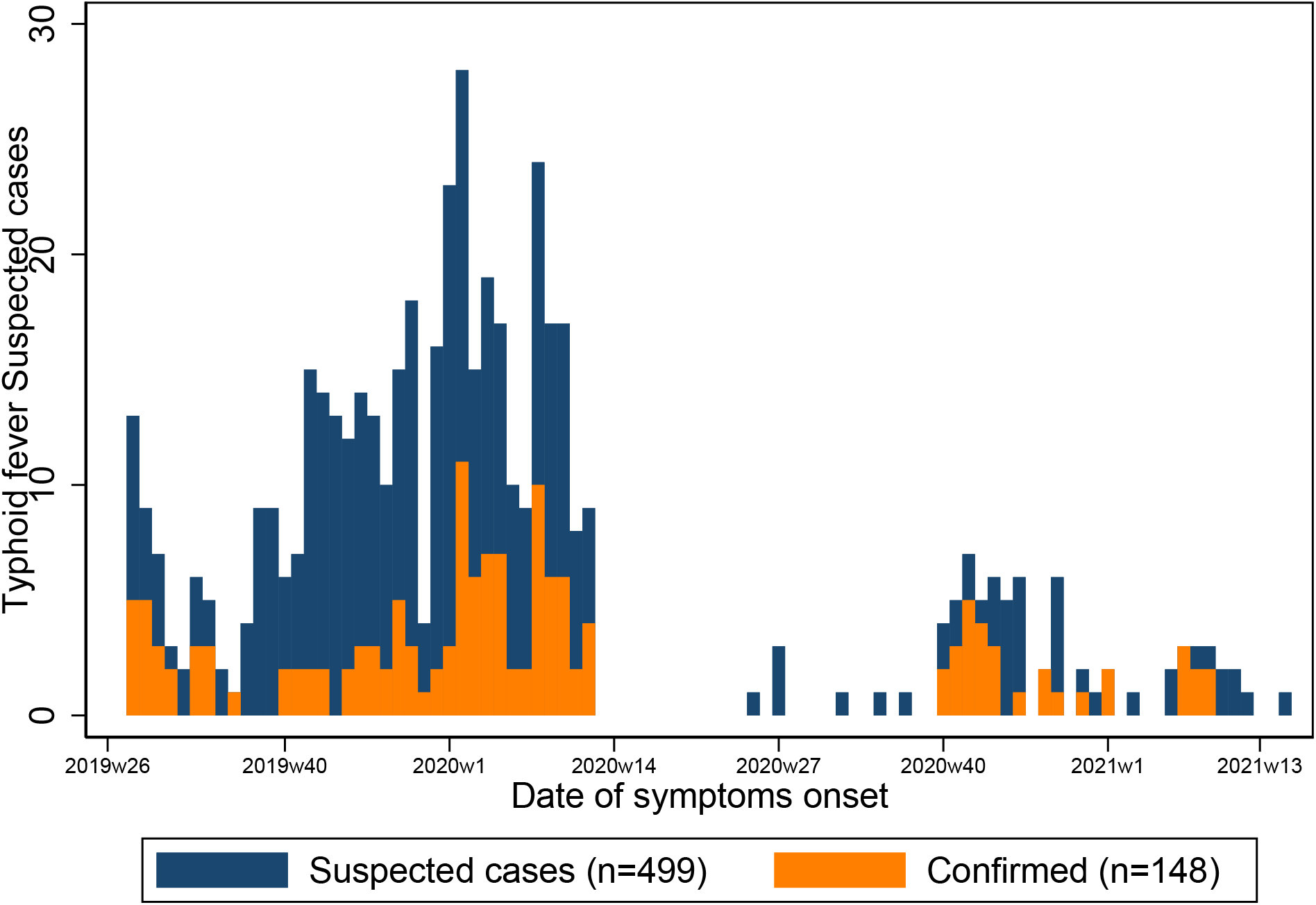
Distribution of suspected typhoid fever cases and confirmed typhoid fever cases by date of symptoms onset over the two study periods, July 2019-March 2020 and July 2021-April 2021 Harare, Zimbabwe

### Descriptive analysis

Among all of the 499 suspected TF cases, 49.1% (n=245) were female and 48.5% (n=242) were aged ≤ 15 years.

The most frequent symptoms among confirmed TF cases were chills, malaise, headache followed by abdominal pain and watery diarrhoea. Among suspected TF cases, 85 (17%) reported starting antibiotics after consulting at the health facility. A tenth of the patients were admitted as inpatients (Supplementary Material 1).

Comparisons of the characteristics of matched TF cases and controls included in the VE analysis are shown in Tables 2 and 3 where we can see the significance differences between facility controls, community controls and cases and probable cases in the primary analysis age group and in the secondary analysis (extended age group) respectively.

**Table 2.** Comparison of characteristics of matched cases and controls included in the effectiveness analysis aged 6 months to 15 years, Harare, Zimbabwe, July 2019 - April 2021.

| Characteristic |  | Matched confirmed cases |  | Matched facility controls |  | Matched p-value | Matched community controls |  | Matched p-value | Matched probable cases |  | Matched facility controls |  | Matched p-value | Matched community controls |  | Matched p-value |
| --- | --- | --- | --- | --- | --- | --- | --- | --- | --- | --- | --- | --- | --- | --- | --- | --- | --- |
|  | Total | 47 |  | 47 |  |  | 224 |  |  | 26 |  | 26 |  |  | 125 |  |  |
|  |  | N | (% ) | N | (% ) |  | N | (% ) |  | N | (% ) | N | (% ) |  | N | (% ) |  |
| Gender | Male | 24 | (51) | 24 | (51) | Matched | 109 | (50) | Matched | 12 | (46) | 12 | (46) | Matched | 65 | (53) | Matched |
|  | Female | 23 | (49) | 23 | (49) | variable | 115 | (50) | variable | 14 | (54) | 14 | (54) | variable | 60 | (47) | variable |
| Age | Mean (SD) | 8.7 | 3.80 | 6.97 | 4.15 | Matched variable | 8.5 | 3.52 | Matched variable | 5.8 | 3.53 | 3.7 | 2.76 | Matched variable | 6.5 | (4) | Matched variable |
| Residential Area | Glen View | 32 | (68) | 32 | (68) | Matched | 149 | (67) | Matched | 10 | (38) | 10 | (38) | Matched | 46 | (37) | Matched |
|  | Mbare | 11 | (23) | 11 | (23) | variable | 55 | (25) | variable | 7 | (27) | 7 | (27) | variable | 36 | (29) | variable |
|  | Kuwadzana | 4 | (9) | 4 | (9) |  | 20 | (9) |  | 9 | (35) | 9 | (35) |  | 43 | (34) |  |
| Years residing in Harare | < 5 years | 10 | (21) | 21 | (45) | 0.004 | 55 | (26) | 0.357 | 13 | (50) | 19 | (73) | 0.019 | 60 | (50) | 0.732 |
|  | 5 to 10 years | 18 | (38) | 13 | (28) |  | 80 | (38) |  | 8 | (31) | 7 | (27) |  | 32 | (27) |  |
|  | > 10 years | 19 | (40) | 13 | (28) |  | 77 | (36) |  | 5 | (19) | 0 | 0 |  | 27 | (23) |  |
|  | *Do not know (excluded)* | 0 |  | 0 |  |  | 12 |  |  | 0 |  | 0 |  |  | 5 |  |  |
| Vaccinated in TCV vaccination campaign (reported) | No | 17 | (36) | 10 | (21) | 0.104 | 35 | (16) | 0.001 | 2 | (8) | 5 | (21) | 0.409 | 26 | (21) | 0.091 |
|  | Yes | 30 | (64) | 37 | (79) |  | 188 | (84) |  | 22 | (92) | 19 | (79) |  | 98 | (79) |  |
|  | *Do not know (excluded)* | 0 |  | 0 |  |  | 1 |  |  | 2 |  | 2 |  |  | 1 |  |  |
| Highest education level achieved | Child < 4 years | 9 | (19) | 17 | (36) | 0.004 | 37 | (17) | 0.068 | 13 | (50) | 17 | (62) | 0.220 | 54 | (43) |  |
|  | None or Primary school | 31 | (66) | 23 | (49) |  | 171 | (76) |  | 13 | (50) | 8 | (35) |  | 67 | (54) |  |
|  | Secondary school | 7 | (15) | 7 | (15) |  | 16 | (7) |  | 0 |  | 1 | (4) |  | 4 | (3) |  |
| Household wealth quintile* | Lowest quintile | 4 | (9) | 5 | (11) | 0.149 | 38 | (17) | 0.075 | 8 | (31) | 5 | (19) | 0.044 | 28 | (23) | 0.018 |
|  | Second quintile | 4 | (9) | 12 | (26) |  | 40 | (18) |  | 1 | (4) | 3 | (12) |  | 35 | (28) |  |
|  | Middle quintile | 11 | (23) | 10 | (21) |  | 57 | (25) |  | 3 | (12) | 11 | (42) |  | 22 | (18) |  |
|  | Fourth quintile | 27 | (58) | 19 | (40) |  | 81 | (36) |  | 14 | (54) | 7 | (27) |  | 39 | (31) |  |
|  | Top quintile | 1 | (2) | 1 | (2) |  | 8 | (4) |  | 0 |  | 0 |  |  | 1 |  |  |
| Household size | Mean (range) | 5.3 | 2 to 15 | 5.0 | 2 to 13 |  | 5.4 | 2 to 16 |  | 4.5 | 3 to 9 | 4.8 | 3 to 8 |  | 5.0 | 2 to 12 |  |
| > 5 persons in the household | No | 30 | (64) | 29 | (62) | 0.835 | 145 | (65) | 0.928 | 23 | (88) | 19 | (73) | 0.199 | 86 | (69) | 0.079 |
|  | Yes | 17 | (36) | 18 | (38) |  | 79 | (35) |  | 3 | (12) | 7 | (27) |  | 39 | (31) |  |
| < 5 year-old in the household | No | 20 | (43) | 16 | (34) | 0.370 | 79 | (35) | 0.457 | 7 | (27) | 4 | (15) | 0.249 | 22 | (18) | 0.321 |
|  | Yes | 27 | (58) | 31 | (66) |  | 145 | (65) |  | 19 | (73) | 22 | (85) |  | 103 | (82) |  |
| Health seeking behaviour: first place to look for care | Health center | 42 | (89) | 46 | (98) | 0.019 | 221 | (99) | 0.004 | 26 | (100) | 26 | (100) |  | 123 | (98) |  |
|  | Others (Private Clinic, pharmacy, others) | 5 | (11) | 1 | (2) |  | 3 | (1) |  |  |  |  |  |  | 2 | (2) |  |
| Mean of transport and time (minutes) | By foot ≤ 20 min | 24 | (51) | 20 | (43) | 0.761 | 119 | (53) | 0.088 | 10 | (38) | 8 | (31) | 0.673 | 74 | (59) | 0.005 |
|  | By foot > 20 min | 14 | (30) | 16 | (34) |  | 76 | (34) |  | 7 | (27) | 8 | (31) |  | 32 | (26) |  |
| to 1<sup>st</sup> place for care | Motor transport (any time) | 6 | (13) | 8 | (17) |  | 26 | (12) |  | 6 | (23) | 5 | (19) |  | 19 | (15) |  |
|  | Unknown transport > 20 min | 3 | (6) | 3 | (6) |  | 3 | (1) |  | 3 | (12) | 5 | (19) |  |  |  |  |
| Contact with a typhoid case | No | 18 | (69) | 19 | (40) | 0.845 | 70 | (91) | 0.195 | 16 | (84) | 15 | (100) |  | 65 | (92) | 0.013 |
|  | Yes, outside the household # | 5 | (19) | 3 | (6) |  | 4 | (5) |  | 1 | (5) | 0 |  |  | 2 | (3) |  |
| | Yes, within the household $\pi$ | 3 | (12) | 3 | (6) | | 3 | (4) | | 2 | (11) | 0 | | | 4 | (6) | |
|  | *Do not know (excluded)* | 21 |  | 22 |  |  | 147 |  |  | 7 |  | 11 |  |  | 54 |  |  |
| Main source of drinking water | Borehole/piped treated | 7 | (15) | 8 | (17) | 0.358 | 56 | (25) | **0.035** | 3 | (12) | 4 | (15) | 0.159 | 26 | (21) | **0.004** |
|  | Borehole untreated | 32 | (68) | 26 | (55) |  | 110 | (49) |  | 22 | (85) | 17 | (65) |  | 70 | (56) |  |
|  | Piped untreated | 5 | (11) | 5 | (11) |  | 25 | (11) |  | 0 | (0) | 3 | (12) |  | 21 | (17) |  |
|  | Other source treated/untreated ^ | 3 | (6) | 8 | (17) |  | 33 | (14) |  | 1 | (4) | 2 | (8) |  | 7 | (6) |  |
| Storing drinking water covered in the household | Yes | 47 | (100) | 47 | (100) |  | 217 | (97) | 0.110 | 26 | (100) | 26 | (100) |  | 123 | (98) |  |
|  | No |  |  |  |  |  | 7 | (3) |  |  |  |  |  |  | 2 | (2) |  |
| Consuming food or drink from the market/street vendor recently | Not at all | 22 | (47) | 27 | (59) | 0.549 | 124 | (56) | **0.041** | 11 | (42) | 12 | (46) | 0.942 | 85 | (68) | **<0.001** |
|  | At least once | 19 | (40) | 14 | (30) |  | 92 | (41) |  | 9 | (35) | 8 | (31) |  | 34 | (27) |  |
|  | Everyday | 6 | (13) | 5 | (11) |  | 7 | (3) |  | 4 | (15) | 6 | (23) |  | 6 | (5) |  |
|  | *Do not know (excluded)* |  |  | 1 |  |  | 1 |  |  | 2 |  | 8 |  |  | 0 |  |  |
| Soap for hand washing in the household | No | 21 | (45) | 18 | (38) | 0.403 | 153 | (68) | **0.001** | 5 | (19) | 12 | (46) | 0.065 | 50 | (40) | **0.049** |
|  | Yes | 26 | (55) | 29 | (62) |  | 71 | (32) |  | 21 | (81) | 14 | (54) |  | 75 | (60) |  |
| Hand washing - Critical moments $\mu$ | ≤ 2 | 12 | (26) | 19 | (40) | 0.226 | 63 | (28) | 0.904 | 10 | (15) | 8 | (15) | 0.554 | 42 | (34) | 0.567 |
|  | 3 | 31 | (66) | 25 | (53) |  | 146 | (65) |  | 14 | (77) | 17 | (81) |  | 78 | (62) |  |
|  | ≥ 4 | 4 | (9) | 3 | (6) |  | 15 | (7) |  | 2 | (8) | 1 | (4) |  | 5 | (4) |  |
| Places used to defecate | Toilet inside the house | 24 | (51) | 26 | (55) | 0.211 | 64 | (29) | **0.014** | 15 | (58) | 10 | (38) | 0.272 | 68 | (55) | 0.112 |
|  | Locked latrine (few families) | 11 | (23) | 8 | (17) |  | 74 | (33) |  | 3 | (12) | 5 | (19) |  | 32 | (25) |  |
|  | Latrine - common use (several families) | 11 | (23) | 8 | (17) |  | 77 | (34) |  | 6 | (23) | 10 | (38) |  | 24 | (19) |  |
|  | Other | 1 | (2) | 5 | (11) |  | 9 | (4) |  | 2 | (8) | 1 | (4) |  | 1 | (1) |  |
| Presence of toilet/latrine in the house or property | Yes, not flooded | 39 | (83) | 42 | (89) | 0.363 | 211 | (94) | **0.005** | 18 | (69) | 20 | (80) | 0.165 | 118 | (95) | **<0.001** |
|  | No, or flooded | 8 | (17) | 5 | (11) |  | 13 | (6) |  | 8 | (31) | 5 | (20) |  | 6 | (5) |  |
| HIV test and status |  |  |  |  |  |  |  |  |  |  |  |  |  |  |  |  |  |
|  | Test not done | 34 | (72) | 32 | (68) | 0.867 | 190 | (85) | **0.021** | 21 | (81) | 22 | (85) | 0.705 | 113 | (91) | 0.231 |
|  | Test done, HIV-negative | 12 | (26) | 14 | (30) |  | 34 | (15) |  | 5 | (19) | 4 | (15) |  | 12 | (9) |  |
|  | Test done, HIV-positive | 1 | (2) | 1 | (2) |  | 0 | 0 |  |  |  |  |  |  | 0 |  |  |

\* estimated based on the presence of assets in the household (radio, motorbike, TV, generator, fridge, oven, mobile phone) # Contact in the 2 previous weeks: within the neighbours or in a funeral or within those sharing latrines or water source $\pi$ Current or during the 2 previous weeks ^ Others include: protected and unprotected shallow wells, surface water, water truck.
Unfrequently: at least once a week $\mu$ Critical moments asked include: before eating, after eating, after going to the toilet, before cooking, after taking care of a child after toilet
For the analysis with community controls, 46 confirmed cases and 27 probable cases were matched

**Table 3.**
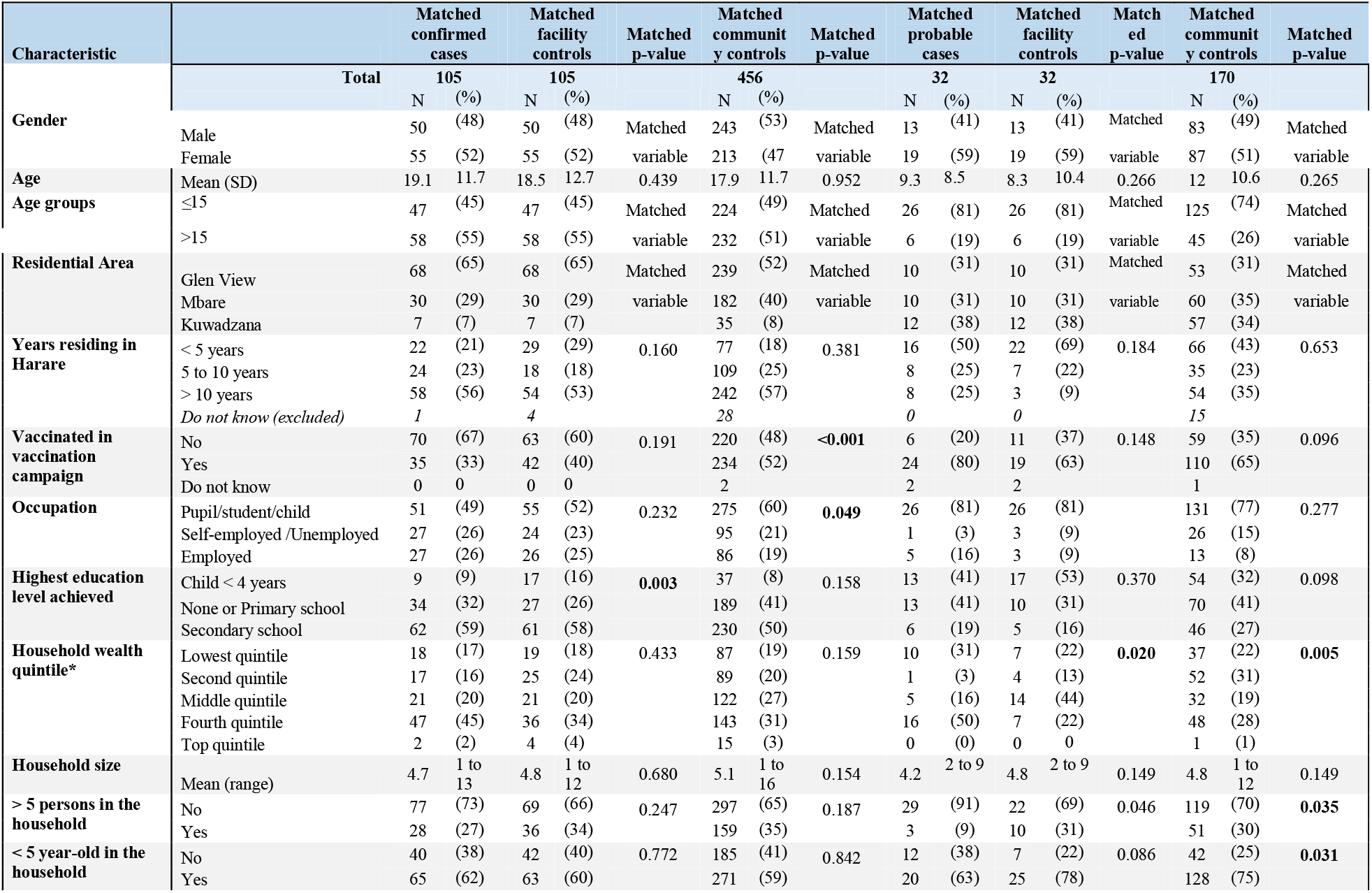
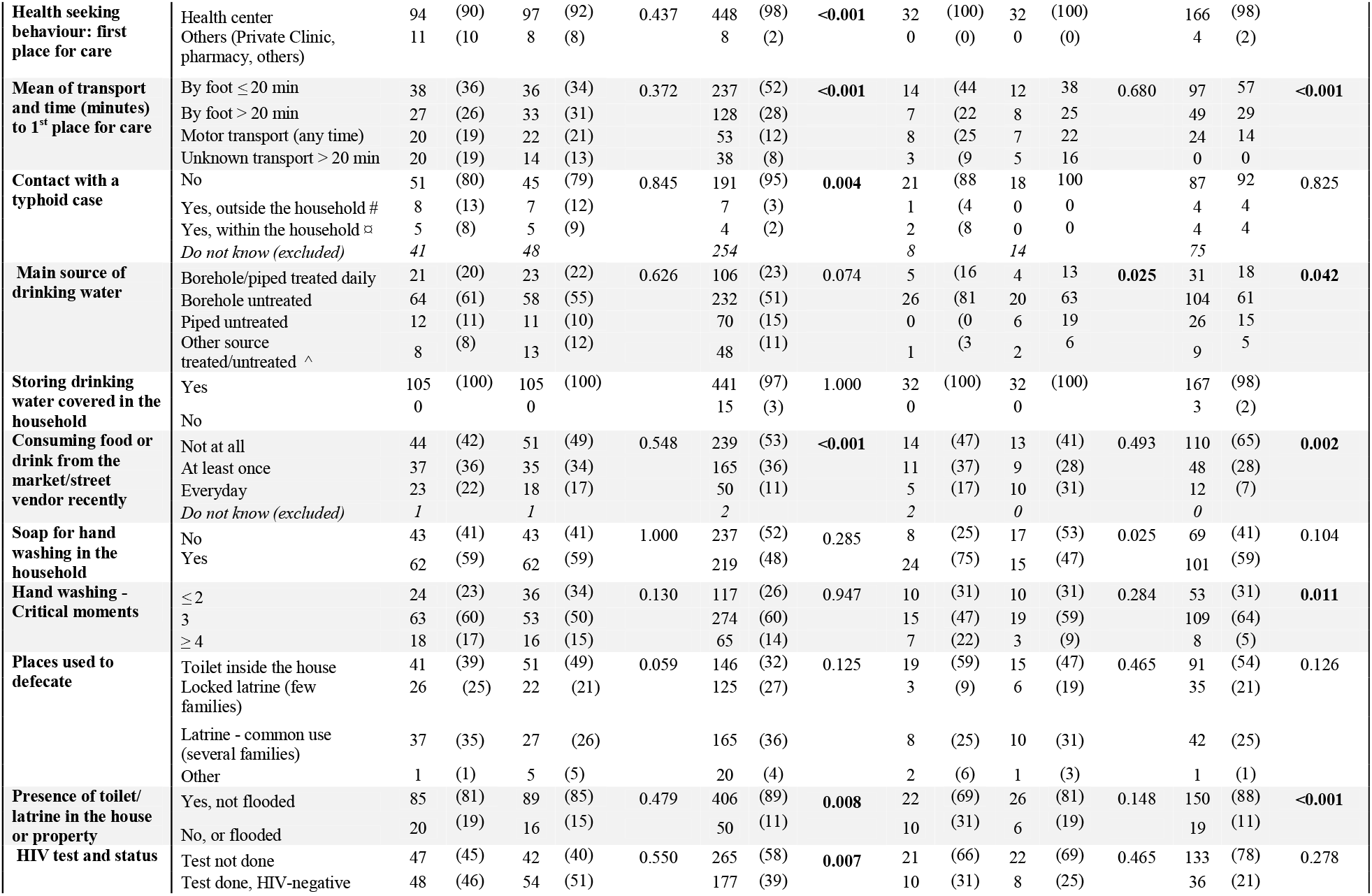
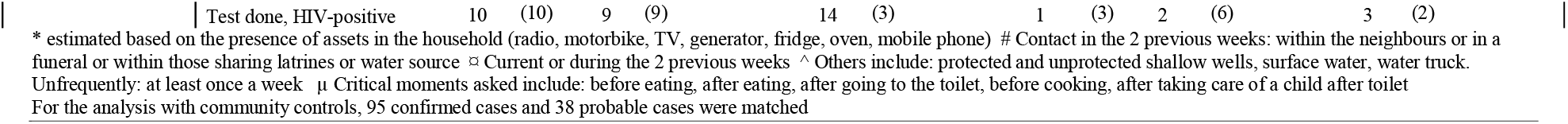
Comparison of characteristics of matched cases and controls included in the effectiveness analysis aged 6 months to 45 years, Harare-Zimbabwe, July 2019 -April 2021.

Several variables were statistically associated with the outcome and the vaccination status and were identified as potential confounders (Supplementary Material 2 and 3).

### Vaccine effectiveness

Table 4 shows the crude and adjusted VE estimates. The adjusted VE against confirmed TF was 75% (95% CI: 1–94, p=0.049) when compared to facility controls, and 84% (95% CI 57–94, p<0.001) when compared to community controls among individuals six months to 15 years old (primary analysis). The adjusted VE against confirmed TF was 46% (95% CI: -26–77, p=0.153) when compared to facility controls, and 67% (95% CI: 35–83, p=0.002) when compared to community controls among individuals six months to 45 years old (secondary analysis). Adjusted VE estimates using probable TF cases were lower in both facility and matched controls.

**Table 4.**
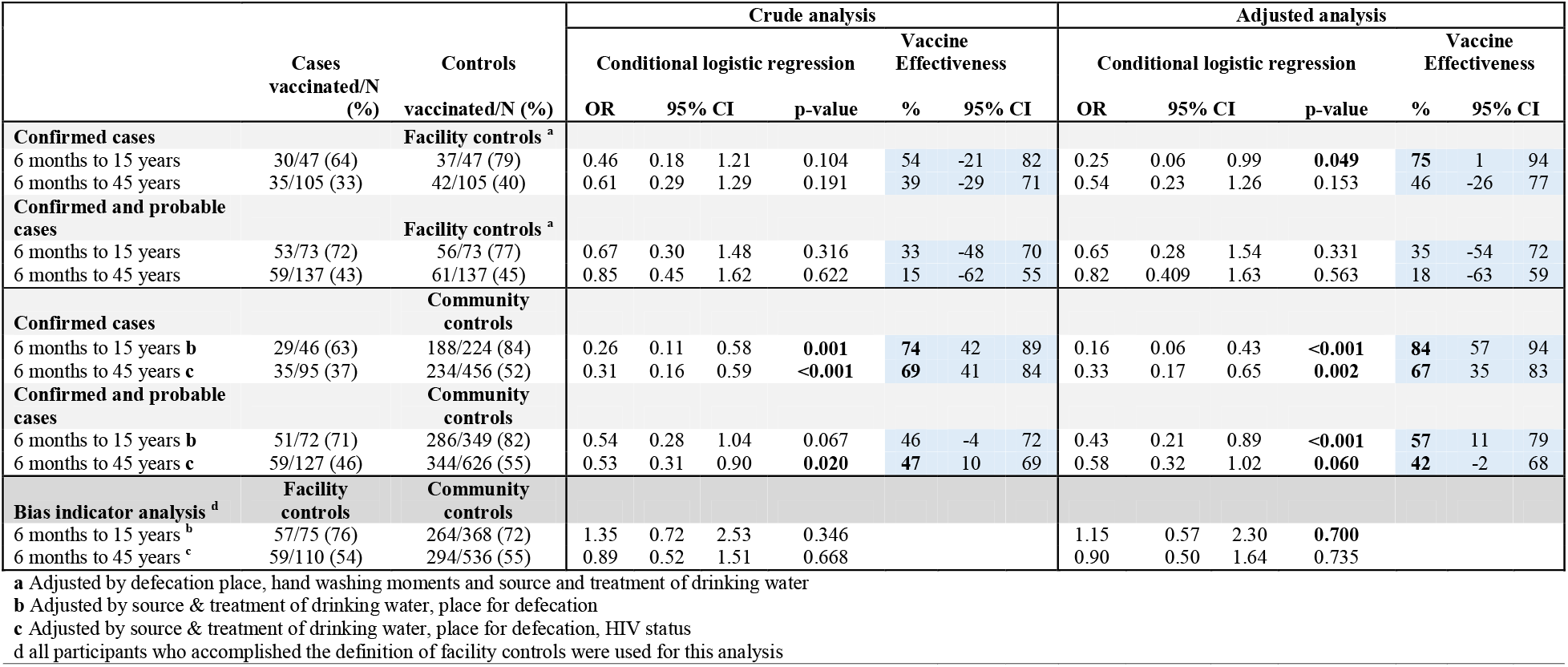
Estimated TCV vaccine effectiveness, crude and adjusted analysis; and indicator bias crude and adjusted analysis.

### Indicator bias analysis and carriage status

When comparing facility controls to matched community controls, there was no difference in the odds of vaccination in the group six months to 15 years (crude OR=1.35, 95%CI 0.72-2.53; adjusted OR=1.15, 95%CI 0.57-2.30), or in the group six months to 45 years (crude OR=0.89, 95%CI 0.52-1.51; adjusted OR=0.90, 95%CI 0.50-1.64, p= 0.735).

One confirmed TF case (1/89, 1.1%) was a convalescent carrier, and there were no (0/55) chronic carriers. Asymptomatic carriage was identified in three (3/1280; 0.23%) community controls.

## Discussion

This study provides the first evidence of VE in an African urban setting where the TCV vaccine was used in a mass campaign in response to an outbreak under real field and programmatic conditions. The results show a significant protective effect of TCV against blood culture-confirmed TF among cases six months to 15 years, but the effect was lower in the secondary analyses (including adults and using probable cases).

These differences might be attributed to the possible naturally induced immunity conferring partial protection among older individuals in the general population (adults in endemic area such as Harare being repeatedly exposed to TF) combined with lower vaccination rates among this group of previously infected and partially protected adults. The mass vaccination campaign targeted adults (>15 years) only in Mbare suburb. A survey conducted in April 2019 in the nine suburbs targeted for the TCV campaign reported an overall coverage of 85% (95% CI: 82– 88) of the TCV vaccine among children six months -15 years and 65% (95% CI 55–73) among adults in Mbare.^16^

In relation to the lower VE estimates obtained for probable TF cases, it is important to note that they were selected from suspected cases after testing negative by blood culture but were positive by sero-diagnosis, *i*.*e*., a duplication of antibody titres between the day of recruitment and day 30. Differential misclassification of the case status among vaccinated and unvaccinated individuals could have occurred (since initial titres and immune response could differ in these two groups)^20^ and lead to underestimation of the VE if variations of titres were higher in vaccinated individuals.

Our VE results are coherent with the available evidence on TCV protection. A challenge study conducted in the UK, showed protective efficacy of 54.6% (95% CI 26.8–71.8) among “naïve” adults (from a non-endemic area) exposed to a large bacterial inoculum. The TF definition potentially included self-limiting asymptomatic bacteraemia and, when restricted to cases who were *S*. Typhi blood-culture-positive, the reported vaccine efficacy was 87.5%.^21^

Furthermore, our results are consistent with previous studies conducted in Asia. One study in Vietnam^22^ reported a vaccine efficacy of 91.5% (95% CI, 77.1-96.6) with two doses of the TCV Vi-rEPA. A phase three trial conducted in India, reported a sero-efficacy estimate of 85% (95% CI, 80– 88) with Vi-TT.^23^ Interim results from an ongoing efficacy trial in Nepal showed a vaccine efficacy of 81.6% (95% CI, 58.8–91.8)^24^ one year after TCV administration among individuals nine months to 16 years old. In a similar age population, a recent cluster randomized clinical trial in Bangladesh showed similar protection provided by Vi-TT (85%; 95% CI 76–91).^25^

During an emergent extensively drug-resistant (XDR) *S*. typhi outbreak in Hyderabad Pakistan, a population-based prospective cohort study estimated VE TF at 95% (95% CI 93–96).^26^ However, in a similar design to our study in a peri-urban community of Karachi, in a matched case-control study following a Vi-TT mass immunisation campaign, the estimated VE was 72% (95% CI 34-88) among children six months to 15 years, which was closer to our results among children in Harare.

In Africa, recent results of a phase three, double blind Vi-TT efficacy trial conducted in Malawi among children nine months to 12 years showed 80.7% protection (95% CI 64.2–89.6) in intention to treat and 83.7% (95% CI 68.1–91.6) in per protocol analysis.^27^

This study is important as it confirms the efficacy of mass vaccination for TF when used under real field and programmatic conditions in the African context as part of an outbreak response.

Data were collected prospectively, using a standardised questionnaire by trained interviewers, with high completeness. During the recruitment in the health facilities, the interviewer and the patients ignored if the TF diagnosis was confirmed, probable or negative (facility controls). So, interviewer and recall bias (*e*.*g*., patients diagnosed with TF would consider potential sources whereas the comparison group would not) although possible are unlikely.

Our control groups appeared representative of the population targeted by the TCV vaccination campaign. Indeed, the vaccine coverage of the six months to 15 years old community controls (82% to 84%) was consistent with the vaccination coverage found in the same age group in a population base survey (85%, 95% CI: 82– 88). Although vaccine coverage tended to be a bit lower among facility controls (77% to 79%), it was consistent with the survey findings. Also, cases were likely representative of cases presenting in health facilities considering there was systematic screening of all TF suspected cases who arrived at the health centres. In addition, the bias indicator analysis showed that the VE estimates unlikely resulted from differential health seeking behaviour between cases and controls.

However, our observational design has limitations. Blood culture was used for the confirmation of the outcome, but its sensitivity is around 60%. The use of serology and of a sensitive classification of probable cases limited the risk of misclassification of TF cases not detected by blood culture.

To confirm vaccination status, interviewers reminded study participants regarding the TCV vaccination campaign, including information about the organisers, dates, and sites, and systematically showed pictures of a TCV vaccine vial. They also tried to confirm TCV vaccination with their vaccination card at home. However, very few vaccinated participants, either cases or controls, could show the vaccination card (many children were vaccinated at school where the vaccination cards were kept). In the absence of a vaccination card, participants were asked to clarify the dates, place, and administration route to confirm that they had received the TCV and not another vaccine (especially the oral cholera vaccine provided in a campaign one month before and after the TCV campaign). Misclassification cannot be excluded but would be likely non-differential (suspected cases were unaware of their diagnosis at the time of the interview).

Information on TF risk factors were obtained through standardised questionnaires, but socially approved behaviours are usually self-reported more frequently than observed. For example, answers to the questions related to hand hygiene and water treatment may have been different in participants interviewed at home (where visual observations would have been possible) or in health centres.

Another limitation of our study is the interruption of the recruitment for some months due to the COVID-19 pandemic. Recruitment of cases and controls finished in April 2021, more than a year after the vaccination campaign (March 2019). However, a study in India showed that 100% of Typbar-TCV recipients across all age groups achieved and maintained sero-protective titers at least two years after vaccination. Our results show that protection was high over a 26-month study period.

Further studies should address the duration of protection conferred by the TCV. Additionally, a better understanding of the full public health impact of the introduction of the vaccine is required, including the possible impact in reducing the spread of multidrug-resistant strains that significantly increase typhoid case fatality risk.

In conclusion, this study confirms that one dose of TCV is an effective tool to prevent TF symptomatic infection among children aged six months to 15 years old. It is the first study assessing the use of the TCV under real programmatic conditions following a mass vaccination campaign in an African setting. These findings are in line with WHO recommendations and the decision made by several countries to introduce TCV into routine vaccination.

### Contributors

PM, FN, MVH, I Panunzi, DG, CD, KM, FJL conceptualised and designed the study. MSL, KK, and FM supervised and monitored data collection. PM, KK, and FM contributed to field operations, data collection, coordination with stakeholders. EF collaborated on site study launching and laboratory serology test selection. AT supervised the laboratory processing. MSL did the statistical analysis and FJL verified the underlying data. MSL prepared the first draft of the manuscript

FN, MVH and FJL substantially reviewed and edited the manuscript. All authors contributed to the interpretation of data, critically reviewed the manuscript, and decided to publish the paper.

### Declaration of interests

We declare no competing interests.

### Data sharing

Individual pseudonymised participant data that underlie the results reported in this article (text, tables, figures, and appendices, with data dictionary) will be made available to others upon submission of a proposal. Requests will be reviewed and sharing of the data will follow the conditions required by all applicable laws and the possible prior signature of any necessary agreement, in accordance with the legal framework set forth by Médecins Sans Frontières (MSF) data sharing policy, which ensures that all security, legal, and ethical concerns are addressed. (https://www.msf.org/sites/msf.org/files/msf_data_sharing_policycontact_infoannexes_final.pdf). For data access and additional related documents such as the study protocol readers may contact Robert Nsaibirni, Data Protection and Compliance Officer.

## Data Availability

Individual pseudonymised participant data that underlie the results reported in this article (text, tables, figures, and appendices, with data dictionary) will be made available to others upon submission of a proposal. Requests will be reviewed and sharing of the data will follow the conditions required by all applicable laws and the possible prior signature of any necessary agreement, in accordance with the legal framework set forth by Medecins Sans Frontieres (MSF) data sharing policy, which ensures that all security, legal, and ethical concerns are addressed.

## Acknowledgments

The study was funded by Médecins Sans Frontières. Epicentre receives core funding from Médecins Sans Frontières. We wish to thank study participants and the team from the Mission Zimbabwe (nurses, GIS officer, wash, logistics and support staff). We also thank Tony Reid from Médecins Sans Frontières for the medical editing support of this manuscript.

All authors attest they meet the ICMJE criteria for authorship.

## Notes

### Competing Interest Statement

The authors have declared no competing interest.

### Funding Statement

The study was funded by Medecins Sans Frontieres

### Author Declarations

The study protocol was approved by the Medical Research Zimbabwe Committee (approval number MRCZ/A/2460) and by the Medecins Sans Frontieres Ethics Review Board (approval number 18114).

